# Hematocrit and Progressive Stroke in Single Small Subcortical Infarction: Insights from the China Stroke Registry III

**DOI:** 10.1101/2024.11.26.24318033

**Authors:** Jiancong Lu, Graeme J. Hankey, Xiaoli Zhang, Xia Meng, Weiqi Zeng, Jun Jiang, Anxin Wang, Zefeng Tan

**Author notes:** Corresponding author, Zefeng Tan, MD,PHD, Address : First People’s Hospital of Foshan, 81 Linnan North Road, Chancheng, Foshan, Guangdong 528000, China. Co-Corresponding author, Anxin Wang. Co-Corresponding author, Jun Jiang.

## Abstract

**Background and Purpose:** Single Small Subcortical Infarction (SSSI) is a subtype of ischemic stroke with unique pathophysiology, often leading to poor outcomes. This study investigates the association between admission hematocrit (HCT) levels and the risk of progressive stroke in SSSI patients.

**Methods:** We conducted a retrospective cohort study using data from the China National Stroke Registry III, including 2,457 SSSI patients. Progressive stroke was defined as an increase in the NIHSS score by ≥2 points during hospitalization. Logistic regression models assessed the association between HCT levels and progressive stroke risk.

**Results:** Among 2,457 SSSI patients, 135 (5.50%) experienced progressive stroke. Higher HCT levels were independently associated with a lower risk of progressive stroke (adjusted OR 0.983, 95% CI 0.973-0.994, P = 0.002), with a potential dose-response relationship. The protective effect was more pronounced in patients with severe sensory deficits (P for interaction = 0.013) and younger age (<65 years) (P for interaction = 0.037). Lower HCT levels were also independently associated with increased mortality risk within 3 months after stroke (adjusted OR 0.937, 95% CI 0.892-0.984, P < 0.001).

**Conclusions:** Higher admission HCT levels are associated with a lower risk of progressive stroke in SSSI patients, particularly those with severe sensory deficits and younger age. Lower HCT levels may also increase early mortality risk. These findings highlight the importance of monitoring and optimizing HCT levels in SSSI management. Further research should explore the underlying mechanisms and therapeutic implications.

## Introduction

Stroke is a leading cause of mortality and disability worldwide, with an estimated 13.7 million new cases and 5.5 million deaths attributed to stroke in 2016.^1^ The global burden of stroke is expected to continue rising due to population growth, aging, and the increasing prevalence of risk factors such as hypertension, diabetes, and obesity.^2^ This growing burden underscores the need for targeted research to improve stroke prevention, treatment, and outcomes. Ischemic stroke accounts for approximately 70% of all stroke cases, with single small subcortical infarction (SSSI), also known as lacunar stroke, representing a distinct subtype that accounts for about 25% of all ischemic strokes.^2,3^

SSSI is a distinct subtype of ischemic stroke characterized by the occlusion of a single perforating artery. Unlike other stroke subtypes that involve larger vessels or multiple occlusions, SSSI affects small, deep brain structures supplied by perforating arteries. This unique pathophysiology may contribute to differences in risk factors, clinical presentations, and outcomes compared to other stroke subtypes.^3^ SSSI is caused by the occlusion of a single perforating artery due to micro atheroma, lip hyalinosis, or perhaps other mechanisms such as endothelial dysfunction and/or blood-brain barrier compromise.^4^ It is characterized by lacunar syndromes like pure motor stroke, pure sensory stroke, sensorimotor stroke, and ataxic hemiparesis. ^5–7^

Although SSSI generally has a more favorable prognosis compared to other stroke subtypes, a significant proportion of patients may experience progressive neurological deterioration during the acute phase, which is associated with poor functional outcomes and increased mortality.^8,9^ The incidence of progressive stroke in SSSI patients varies from 9% to 40% across different studies, depending on the definition and assessment methods used.^8,10^ Identification of predictive factors for progressive stroke in SSSI is crucial for risk stratification and management.

Hematocrit (HCT), the volume percentage of red blood cells in whole blood, is a readily available and routinely measured hematological parameter that reflects the balance between red blood cell production and loss.^10^ Some studies have suggested that high HCT on admission is an independent predictor of poor functional outcome and increased mortality^11–13^, while others have reported that low HCT is associated with larger infarct volume and worse neurological deficits.^14,15^

Despite the growing evidence on the role of HCT in acute ischemic stroke, its association with progressive stroke in SSSI patients remains poorly understood. For instance, Furlan et al. reported that higher admission HCT levels were linked to a 15% lower risk of early neurological deterioration within the first 48 hours.^12^ Given the distinct pathophysiology of SSSI and the lack of specific studies in this population, it is crucial to investigate whether the relationship between HCT and stroke outcomes observed in other stroke subtypes holds true for SSSI. Clarifying this relationship may provide insights into the mechanisms of neurological deterioration in SSSI and guide the development of targeted management strategies.

Therefore, we aimed to investigate the correlation between admission HCT levels and the risk of progressive stroke during hospitalization in a large cohort of SSSI patients from the China National Stroke Registry III (CNSR-III). Although higher HCT could theoretically impair blood flow due to increased viscosity, it may also enhance oxygen delivery to ischemic regions. Supporting this, Furlan et al. reported that higher admission HCT levels were linked to a 15% lower risk of early neurological deterioration within the first 48 hours.^12^ Thus, we hypothesized that lower HCT levels would be independently associated with an increased risk of progressive stroke in SSSI patients.

## Methods

### Study Population

We conducted a retrospective cohort study using data from the CNSR-III (IRB approval number: KY2015-001-01), a nationwide, multicenter, prospective registry of acute ischemic stroke patients in China.^16^ The detailed design and methods of the CNSR-III have been described previously^17^. Briefly, consecutive patients with acute ischemic stroke or transient ischemic attack within 7 days of symptom onset were enrolled from 201 hospitals across China between August 2015 and March 2018. For this study, we included patients with a diagnosis of SSSI confirmed by magnetic resonance imaging (MRI). SSSI was defined as a single subcortical infarct with a maximum diameter ≤15 mm on diffusion-weighted imaging (DWI), located in the territory of the perforating branches of the middle cerebral artery or basilar artery.^5^ The ≤15 mm size criterion for SSSI was based on the traditional definition of lacunar infarcts, which are small, deep infarcts resulting from the occlusion of a single perforating artery.^18^ This size cut-off is thought to reflect the limited territory supplied by a single perforating artery and has been widely used in research on lacunar stroke to distinguish it from larger subcortical infarcts that may have different etiologies and outcomes.^19^

We excluded patients with multiple cerebral infarctions, cortical infarcts, or incomplete clinical data. Additionally, patients without new-onset cerebral infarction within the 7- day time-frame were excluded. Patients with evidence of systemic inflammation or infection on admission, which could affect HCT levels and stroke outcomes, were also excluded. Finally, patients with missing HCT data were excluded from the study to ensure data integrity (Figure 1).

**Figure 1:**
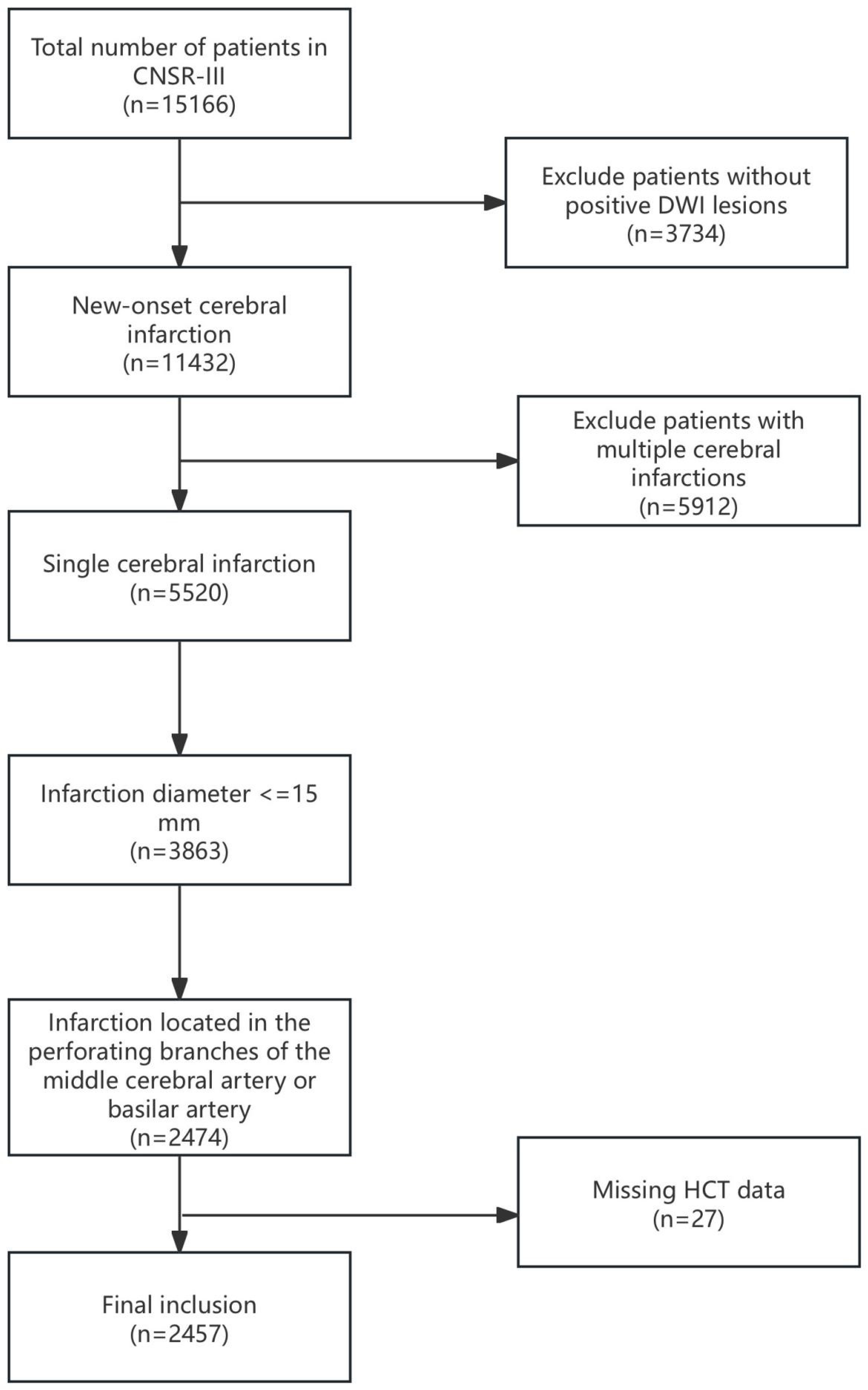
Flowchart of Patient Selection

### Data Collection

Demographic characteristics, vascular risk factors, stroke severity on admission as assessed by the National Institutes of Health Stroke Scale (NIHSS) score, HCT levels, and other laboratory parameters were collected using a standardized case report form. Demographic and clinical data were collected by trained research coordinators using standardized case report forms. The collected data were double-checked by a second coordinator to ensure accuracy and completeness. Any discrepancies were resolved by referring to the original medical records or through discussion with the treating physicians. Neurological status was evaluated daily during hospitalization using the NIHSS score. Functional outcomes were assessed using the modified Rankin Scale (mRS) at 3 months, 6 months and 12 months after stroke.

### MRI Examination and Analysis Protocol

Every subject was scanned using a 3 Tesla MRI system. The imaging protocol included various sequences such as three-dimensional time-of-flight magnetic resonance angiography (MRA) with parameters like a repetition time of 20–25 milliseconds, echo time of 3.3–3.9 milliseconds, a flip angle between 15° and 20°, and a slice thickness ranging from 0.65 to 1.0 mm. Additional sequences performed were axial T2-weighted imaging (with a repetition time of 4500 milliseconds and an echo time of 8 milliseconds), T1-weighted imaging (repetition time of 1200 milliseconds, echo time of 11 milliseconds), fluid-attenuated inversion recovery sequences (repetition time of 7000 milliseconds, echo time of 94 milliseconds), and DWI (repetition time of 3000 milliseconds, echo time of 75 milliseconds). Except for MRA, all these sequences were conducted with a slice thickness of 5 mm and a 1.5 mm gap between slices.^17^

The MRIs were obtained in digital format from different centers and were centrally evaluated by two independent experts (JJ and Y-YX), who were not aware of the patients’ clinical information. In cases of disagreement, they came to a mutual decision.^17^

For the diagnosis of small subcortical strokes on DWI, the criteria included identifying a lesion smaller than 15 mm in diameter. The assessment of intracranial arteries utilized three-dimensional time-of-flight MRA to identify any stenosis in the middle cerebral artery elated to basal ganglia strokes or in the basilar artery related to pontine strokes. The location of infarcts and stenosis was determined to be in the same sagittal or axial plane as appropriate, based on a comprehensive evaluation using various weighted sequences. ^17^

### Outcome Assessment

The primary outcome was an increase in the NIHSS score by ≥2 points during hospitalization, indicating progressive stroke. This threshold was chosen to capture clinically meaningful neurological deterioration in real-time, which could occur at different stages depending on each patient’s condition. Although the duration of hospitalization can vary due to medical and non-medical factors, daily NIHSS assessments allowed for comprehensive monitoring and improved detection of changes in patient status.

Previous studies, such as Furlan et al., have used specific time points like 48 hours to identify early neurological deterioration. ^12^However, we opted for continuous monitoring throughout the hospital stay to better reflect real-world clinical practice. Future studies could explore the utility of evaluating NIHSS changes at standardized time points (e.g., 3, 5, or 7 days) to determine whether specific intervals provide additional insights into the timing and progression of stroke.

The choice of the ≥2-point increase as a threshold was based on its established clinical relevance, providing a clear marker of significant neurological deterioration. Several studies support the use of a ≥2-point increase in NIHSS score as a clinically meaningful indicator of worsening stroke outcomes. ^4^Secondary outcomes included functional dependency, measured by the modified Rankin Scale (mRS) score >2 at 3 months, 6 months, and 12months post-stroke; all-cause mortality at 3 months, 6 months, and 12months post-stroke; and NIHSS worsening by ≥1, ≥3, and ≥4 points during hospitalization. All follow-up assessments and evaluations of outcomes were meticulously conducted in accordance with the CNSR-III protocol guidelines.^17^ These assessments included standardized interviews and examinations at specified time points post-stroke to ensure consistent and accurate data collection.

### Statistical Analysis

Continuous variables were summarized as mean ± standard deviation (SD) for normally distributed data, and as median with interquartile range (IQR) for non-normally distributed data. Group comparisons were performed using the independent samples t-test for normally distributed variables and the Mann-Whitney U test for non-normally distributed variables. Categorical variables were expressed as counts and percentages and were analyzed using the chi-square test or Fisher’s exact test as appropriate.

HCT was analyzed as both a continuous and a categorical variable. For categorical analysis, HCT was divided into quartiles: Q1 (≤38.1%), Q2 (38.2%-41.6%), Q3 (41.7%-44.8%), and Q4 (≥44.9%). Additionally, dichotomous (above vs. below median) and tertile categorizations were used for sensitivity analysis to ensure the robustness of the findings.

Logistic regression models were used to assess the association between HCT levels and the risk of progressive stroke. Models were adjusted sequentially as follows: Model 1 (unadjusted), Model 2 (adjusted for age and sex), and Model 3 (further adjusted for variables that were significant in univariate analyses at P<0.05 and other clinically relevant variables identified from Table 1). Secondary outcomes were analyzed similarly, and subgroup analyses were conducted to explore the effects of HCT on different severities of sensory and motor deficits.

**Table 1:**
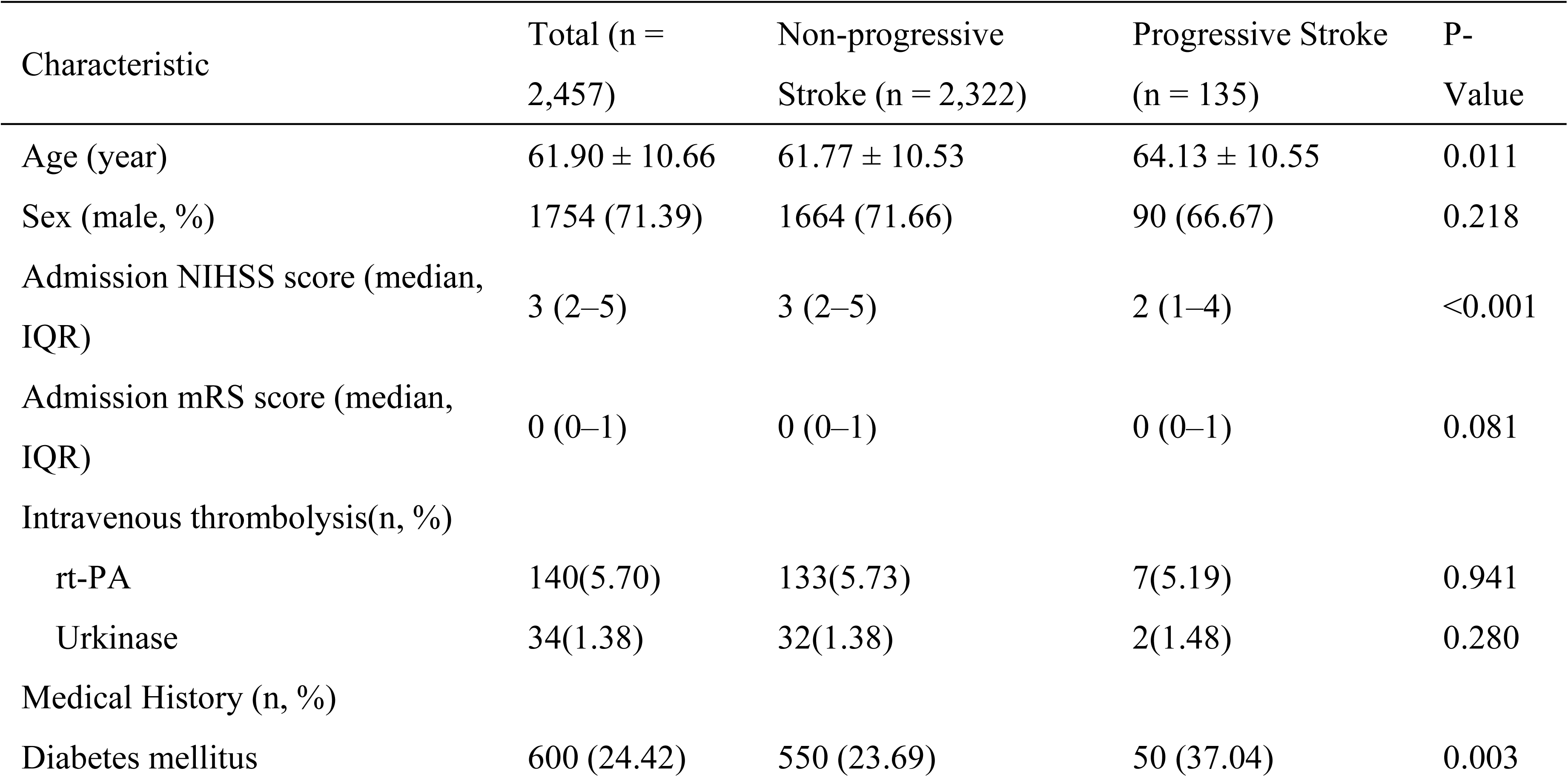
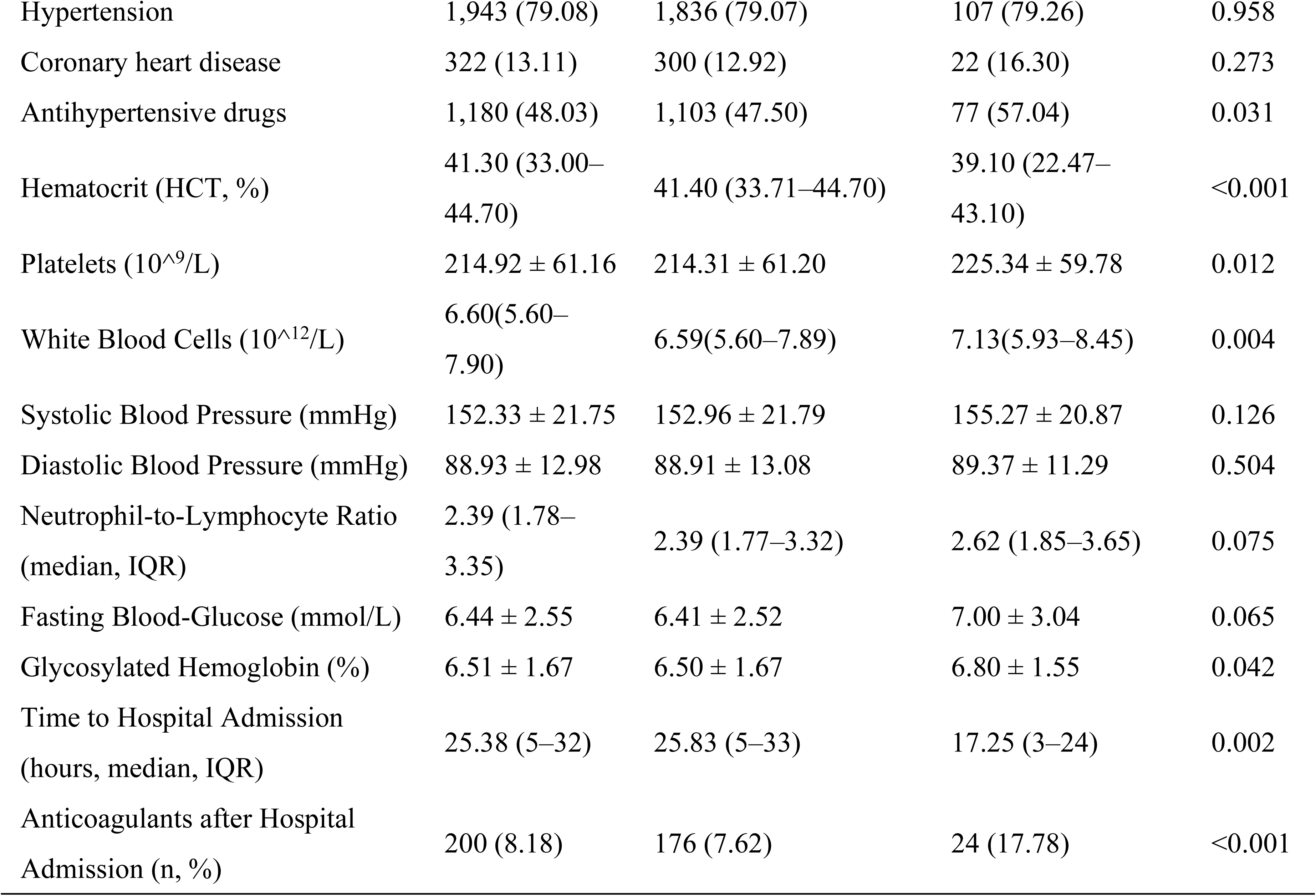
Baseline Characteristics of Patients with Single Small Subcortical Infarction.

| Characteristic | Total (n = 2,457) | Non-progressive Stroke (n = 2,322) | Progressive Stroke (n = 135) | P-Value |
| --- | --- | --- | --- | --- |
| Age (year) | 61.90 ± 10.66 | 61.77 ± 10.53 | 64.13 ± 10.55 | 0.011 |
| Sex (male, %) | 1754 (71.39) | 1664 (71.66) | 90 (66.67) | 0.218 |
| Admission NIHSS score (median, IQR) | 3 (2–5) | 3 (2–5) | 2 (1–4) | <0.001 |
| Admission mRS score (median, IQR) | 0 (0–1) | 0 (0–1) | 0 (0–1) | 0.081 |
| Intravenous thrombolysis(n, %) |  |  |  |  |
| rt-PA | 140(5.70) | 133(5.73) | 7(5.19) | 0.941 |
| Urokinase | 34(1.38) | 32(1.38) | 2(1.48) | 0.280 |
| Medical History (n, %) |  |  |  |  |
| Diabetes mellitus | 600 (24.42) | 550 (23.69) | 50 (37.04) | 0.003 |
| Hypertension | 1,943 (79.08) | 1,836 (79.07) | 107 (79.26) | 0.958 |
| Coronary heart disease | 322 (13.11) | 300 (12.92) | 22 (16.30) | 0.273 |
| Antihypertensive drugs | 1,180 (48.03) | 1,103 (47.50) | 77 (57.04) | 0.031 |
| Hematocrit (HCT, %) | 41.30 (33.00–44.70) | 41.40 (33.71–44.70) | 39.10 (22.47–43.10) | <0.001 |
| Platelets (10<sup>9</sup>/L) | 214.92 ± 61.16 | 214.31 ± 61.20 | 225.34 ± 59.78 | 0.012 |
| White Blood Cells (10<sup>12</sup>/L) | 6.60(5.60–7.90) | 6.59(5.60–7.89) | 7.13(5.93–8.45) | 0.004 |
| Systolic Blood Pressure (mmHg) | 152.33 ± 21.75 | 152.96 ± 21.79 | 155.27 ± 20.87 | 0.126 |
| Diastolic Blood Pressure (mmHg) | 88.93 ± 12.98 | 88.91 ± 13.08 | 89.37 ± 11.29 | 0.504 |
| Neutrophil-to-Lymphocyte Ratio (median, IQR) | 2.39 (1.78–3.35) | 2.39 (1.77–3.32) | 2.62 (1.85–3.65) | 0.075 |
| Fasting Blood-Glucose (mmol/L) | 6.44 ± 2.55 | 6.41 ± 2.52 | 7.00 ± 3.04 | 0.065 |
| Glycosylated Hemoglobin (%) | 6.51 ± 1.67 | 6.50 ± 1.67 | 6.80 ± 1.55 | 0.042 |
| Time to Hospital Admission (hours, median, IQR) | 25.38 (5–32) | 25.83 (5–33) | 17.25 (3–24) | 0.002 |
| Anticoagulants after Hospital Admission (n, %) | 200 (8.18) | 176 (7.62) | 24 (17.78) | <0.001 |

All statistical analyses were conducted using SAS version 9.4 (SAS Institute Inc., Cary, NC, USA). A two-tailed P value <0.05 was considered statistically significant.

## Results

### Baseline Characteristics

Among the 2,457 patients with SSSI, 135 (5.50%) experienced progressive stroke, defined as an increase in NIHSS by ≥2 points during hospitalization. Patients with progressive stroke were older (mean age: 64.13 ± 10.55 years) compared to those without progressive stroke (mean age: 61.77 ± 10.53 years, P = 0.011). Both groups were predominantly male (71.39%), with no significant difference in gender distribution (P = 0.218). The median NIHSS score on admission was significantly higher in the progressive stroke group (P < 0.001). Of the total cohort, 140 patients (5.70%) received recombinant tissue plasminogen activator (rTPA) treatment. There was no statistically significant association between progressive stroke and rTPA treatment status (Chi-square = 0.070, p = 0.792), suggesting that changes in NIHSS scores were not significantly related to rTPA administration in this cohort. (Table1).

Laboratory findings indicated that patients with progressive stroke had significantly lower mean HCT levels (29.66 ± 18.98%) compared to those without progressive stroke (36.12 ± 15.57%; P < 0.001). Additionally, progressive stroke patients had higher platelet counts (mean PLT: 225.34 ± 59.78 vs. 214.31 ± 61.20; P = 0.012), white blood cell counts (median, IQR: 7.13 [5.93–8.45] vs. 6.59 [5.60–7.89]; P = 0.004), and HbA1c levels (mean: 6.80 ± 1.55 vs. 6.50 ± 1.67; P = 0.042).

Progressive stroke patients also had a higher prevalence of diabetes mellitus (37.04% vs. 23.69%; P = 0.003), were more likely to receive antihypertensive drugs prior to admission (57.04% vs. 47.50%; P = 0.031), and more frequently received anticoagulant therapy after admission (17.78% vs. 7.62%; P < 0.001). Notably, the onset-to-admission time was significantly shorter in the progressive stroke group (median, IQR: 17.25 [3– 24] vs. 25.83 [5–33] hours; P = 0.002) (Table 1).

### Association between HCT and Progressive Stroke

Higher HCT levels were significantly associated with a reduced risk of progressive stroke during hospitalization. In the unadjusted logistic regression model, each unit increase in HCT was associated with a 2.1% lower risk of NIHSS worsening by ≥2 points (OR = 0.98, 95% CI, 0.97–0.99; P < 0.001). After adjusting for potential confounders including age, sex, diabetes mellitus, hypertension, baseline NIHSS score, history of prior stroke, and other relevant clinical factors such as platelet count and anticoagulant therapy (Model 3), the association remained significant (adjusted OR = 0.98, 95% CI, 0.97–0.99; P = 0.002). A forest plot (Figure 2) illustrates this association across varying NIHSS thresholds and other clinical endpoints.

**Figure 2:**
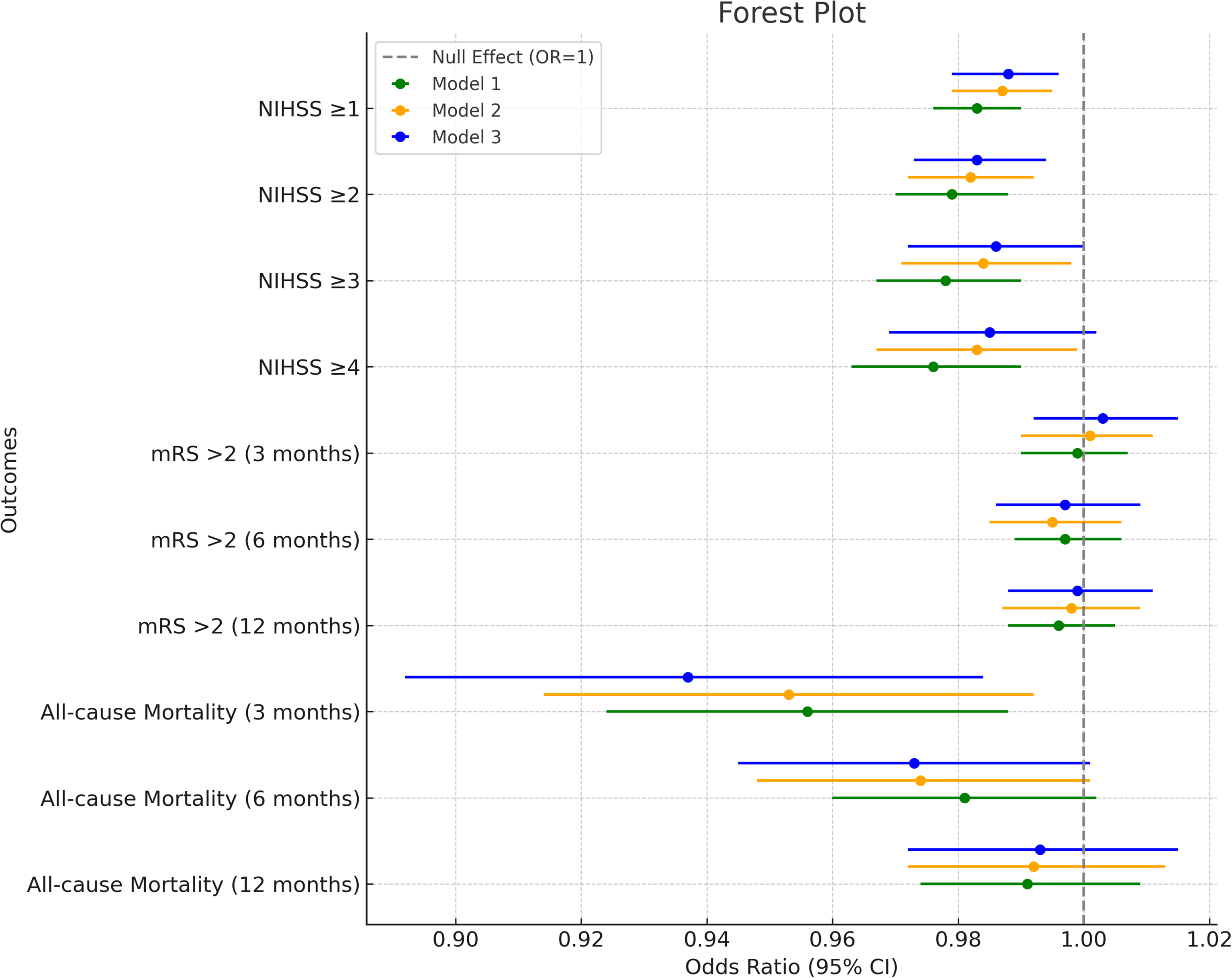
Forest Plot of the Association between Hematocrit (HCT) Levels and Stroke Outcomes

When HCT was analyzed categorically, patients in the highest quartile (≥ 44.9%) had a significantly lower risk of NIHSS worsening by ≥2 points compared to those in the lowest quartile (≤38.1%) (adjusted OR = 0.46, 95% CI, 0.24–0.85; P = 0.013). Similarly, patients with HCT levels above the median (≥41.3%) exhibited a 40.9% reduced risk, though this association did not reach statistical significance (adjusted OR = 0.59, 95% CI, 0.26–1.34; P = 0.209) (Appendix Table 2).

**Table 2:**
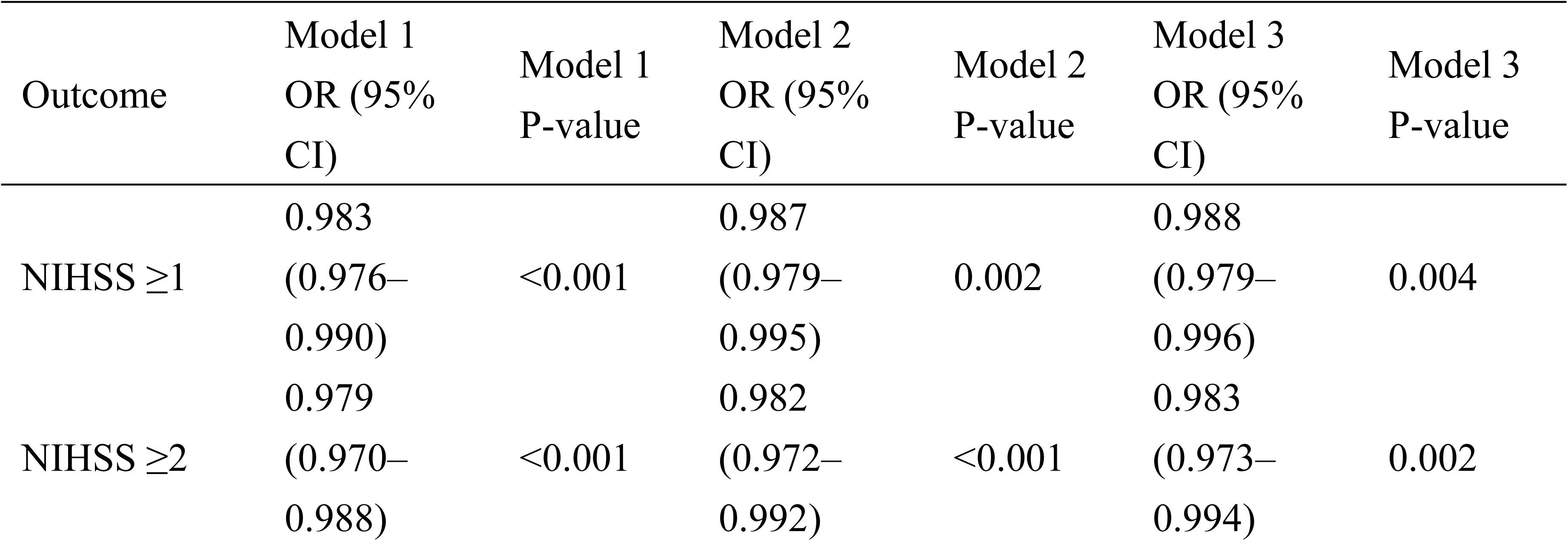
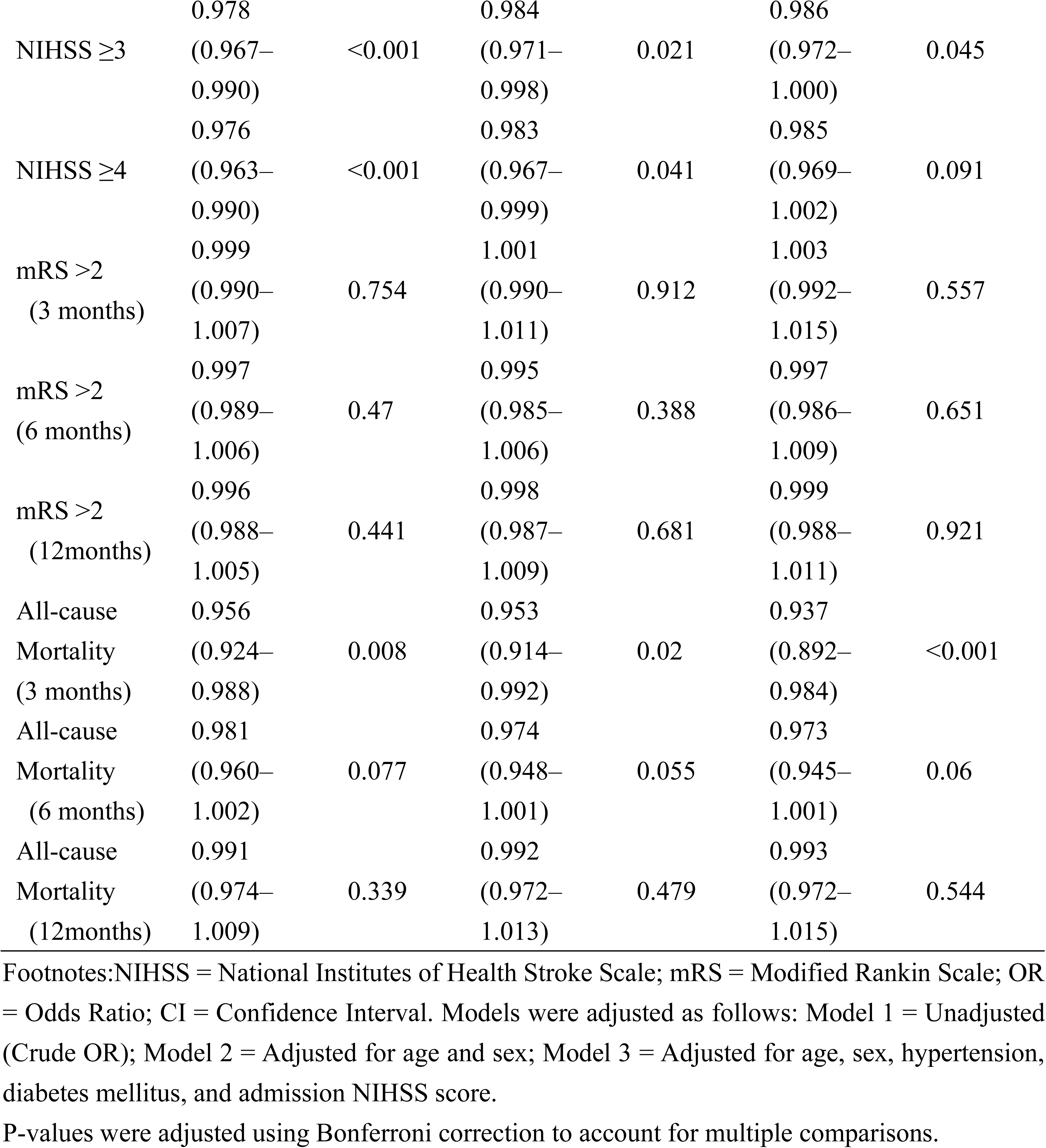
Multivariate Analysis of Progressive Stroke and Mortality Risk.

Furthermore, when using different cut-off points for HCT, patients with levels above 38.4% demonstrated a 32.6% lower risk of NIHSS worsening by ≥2 points. However, this association did not reach statistical significance in the fully adjusted model (adjusted OR = 0.67, 95% CI, 0.32–1.43; P = 0.304). The forest plot of the multivariate regression results for HCT and ΔNIHSS ≥2 points, based on different classification methods, is presented in Appendix Table 2, (Figure 2) .

These findings consistently demonstrate a potential dose-response relationship between higher HCT levels and a reduced risk of progressive stroke in SSSI patients. This protective effect was most pronounced among patients in the highest HCT quartile, underscoring the clinical importance of maintaining HCT levels within an optimal range.

### Subgroup Analyses

Subgroup analyses revealed that the protective effect of higher HCT levels was particularly pronounced in patients with severe sensory or motor deficits. Among patients with a sensory subscale score ≥2 points, each 1% increase in HCT was associated with a 7.0% reduction in the risk of NIHSS worsening by ≥2 points after multivariate adjustment (Model 3, OR = 0.93, 95% CI, 0.88–0.98; P = 0.008) (Appendix Table 7). Similar findings were observed for patients with a sensory subscale score ≥3 points, where higher HCT levels independently predicted a lower risk of NIHSS worsening by ≥3 points (Model 3, OR = 0.93, 95% CI, 0.88–0.98; P = 0.008) (Appendix Table 8). A significant interaction between HCT and sensory deficits for NIHSS was worsed by ≥4 points (P for interaction = 0.013 in the fully adjusted model; Appendix Table 9). This finding indicates that the protective effect of higher HCT levels is more pronounced in patients with severe sensory deficits (subscale score ≥4 points), where each 1% increase in HCT was associated with a 7.3% reduction in the risk of NIHSS worsening by ≥4 points (adjusted OR = 0.93, 95% CI, 0.87–0.99; P = 0.015).

In contrast, the association between HCT and progressive stroke was not statistically significant in patients with mild motor deficits (motor subscale score <1 point) (Appendix Table 5). However, among patients with more severe motor deficits (motor subscale score ≥1 point), higher HCT levels were significantly associated with a lower risk of NIHSS worsening by ≥1 point (Model 3, adjusted OR = 0.92, 95% CI, 0.88– 0.97; P = 0.002) and ≥2 points (adjusted OR = 0.96, 95% CI, 0.93–0.98; P < 0.001) (Appendix Table 5).

These findings suggest that the protective effect of higher HCT levels against progressive stroke may be more pronounced in SSSI patients with severe sensory or motor deficits. The significant interaction between HCT and sensory deficits for NIHSS worsening by ≥4 points further highlights the potential role of sensory dysfunction in modulating the relationship between HCT and progressive stroke severity. These results underscore the importance of accounting for baseline neurological deficits when assessing the impact of HCT on stroke outcomes and may help guide personalized management strategies for SSSI patients.

### Interaction Effects

Interaction analyses revealed significant effects between HCT levels and age, as well as smoking status, on the risk of progressive stroke. A significant interaction was observed between HCT and age on the risk of progressive stroke (ΔNIHSS ≥ 2) (P for interaction = 0.037 in the fully adjusted model). The protective effect of higher HCT levels was more pronounced in younger patients (<65 years) (adjusted OR = 0.97, 95% CI, 0.96–0.99; P < 0.001), compared to older patients (≥65 years) (adjusted OR = 0.99, 95% CI, 0.98–1.01; P = 0.455) (Appendix Table 7). These findings suggest that age may play a critical role in modulating the relationship between HCT and stroke progression.

A borderline significant interaction effect between HCT and smoking status was also observed, suggesting a potentially stronger protective effect in smokers (adjusted OR = 0.97, 95% CI, 0.95–0.99; P < 0.001) compared to non-smokers (adjusted OR = 0.99, 95% CI, 0.98–1.01; P = 0.196) (Appendix Table 7). Further research is needed to confirm and understand these findings.

No significant interactions were observed between HCT and sex, onset-to-admission time, arterial stenosis, platelet parameters, infarct location, or baseline NIHSS score on the risk of progressive stroke (all P for interaction > 0.05). The results for additional interaction effects across varying endpoints (ΔNIHSS ≥ 1, 3, or 4) are presented in Appendix Tables 6, 8, and 9.

### Association between HCT and Other Outcomes

While higher HCT levels were protective against progressive stroke in the acute phase, their association with long-term functional outcomes (mRS > 1 or mRS > 2) at 3 months, 6 months, and 12 months after stroke was not statistically significant in adjusted models (Figure 2). This suggests that the impact of higher HCT levels on long-term functional recovery may be limited or influenced by other factors not accounted for in this study.

In contrast, lower HCT levels were significantly associated with an increased risk of all-cause mortality within 3 months after stroke in both unadjusted (OR = 0.96, 95% CI, 0.92–0.99; P = 0.008) and fully adjusted models (adjusted OR = 0.94, 95% CI, 0.89– 0.98; P < 0.001) (Figure 2). This finding indicates that lower HCT levels may be an independent risk factor for early mortality in SSSI patients, even after adjusting for other relevant variables.

However, the association between HCT and all-cause mortality was not significant at 6 months and 12 months after stroke in both unadjusted and adjusted models (Figure 2), suggesting that the impact of HCT on mortality may be limited to the early post-stroke period, with other factors playing a more prominent role in determining long-term survival.

These findings underscore the complex relationship between HCT levels and various stroke outcomes in SSSI patients, indicating that while higher HCT may offer protection in the acute phase, other factors likely contribute to long-term functional recovery and survival. Future studies with larger sample sizes and extended follow-up periods are needed to clarify the role of HCT in predicting long-term outcomes in SSSI patients. Such research could inform the development of targeted management strategies aimed at optimizing HCT levels to improve both short- and long-term stroke outcomes.

## Discussion

In this large, nationwide cohort study of SSSI patients, we demonstrated that higher admission HCT levels were independently associated with a reduced risk of progressive stroke, defined as an increase in NIHSS by ≥2 points during hospitalization. This protective effect was particularly pronounced in patients with severe sensory deficits, where a significant interaction between HCT and sensory dysfunction was observed for NIHSS worsening by ≥4 points. Additionally, a potential dose-response relationship was identified, with patients in the highest HCT quartile having a significantly lower risk of progressive stroke compared to those in the lowest quartile. However, HCT levels were not significantly associated with functional outcomes at 3 months, 6 months, and 12 months after stroke. In contrast, lower HCT levels were independently associated with an increased risk of all-cause mortality within 3 months post-stroke.

### Association of HCT Levels with Progressive Stroke in SSSI

Our findings are consistent with previous research indicating an inverse association between HCT levels and early neurological deterioration in patients with acute ischemic stroke.^11,12,14,20^ However, this study extends those findings by focusing specifically on SSSI, a stroke subtype with unique pathophysiological characteristics. To our knowledge, this is among the first studies to demonstrate an independent association between higher HCT levels and a lower risk of progressive stroke in SSSI patients, revealing a potential dose-response relationship and a significant interaction effect with sensory deficits.

For instance, in a retrospective study involving 2454 patients with acute ischemic stroke, Furlan et al. reported that higher admission HCT levels were linked to a 15% lower risk of early neurological deterioration within the first 48 hours.^12^ This observation aligns closely with our findings in the SSSI population, where higher HCT levels were associated with reduced risks of progressive stroke.

Furthermore, Kimberly et al. demonstrated a correlation between lower hemoglobin levels and larger stroke volumes, supporting the notion that higher HCT or hemoglobin levels may offer protection against more severe stroke outcomes.^14^ Their study showed a mean stroke volume of 48.6 mL in the lowest hemoglobin quartile versus 37.2 mL in the highest quartile, highlighting the physiological mechanism by which higher HCT could enhance oxygen delivery and mitigate brain tissue damage, consistent with our findings.

Similarly, Sico et al. reported that anemia was associated with poor outcomes in patients with less severe ischemic strokes, with an adjusted odds ratio of 1.3 for poor functional outcomes in anemic patients compared to non-anemic patients.^20^ This reinforces the detrimental impact of low HCT levels on stroke recovery and underscores the importance of maintaining optimal HCT levels to improve prognosis, which aligns with our observations.

Zhang et al. also identified hemoglobin concentration as a key predictor of clinical outcomes in patients with acute ischemic stroke or transient ischemic attack, with those in the lowest hemoglobin quintile showing a significantly higher risk of adverse outcomes.^15^ This finding emphasizes the relevance of blood parameters in stroke prognosis and highlights the importance of HCT monitoring. Additionally, Jeong et al. identified neuroimaging markers for early neurological deterioration in single small subcortical infarction, noting that the presence of these markers was linked to a 2.5- fold increased risk of early deterioration.^8^ Their findings underscore the value of early identification and management of risk factors like HCT. Our study complements this evidence by demonstrating that higher HCT levels are associated with a reduced risk of progressive stroke, suggesting that integrating neuroimaging markers with HCT monitoring could enhance early detection and intervention strategies.

### Mechanisms Linking HCT and Progressive Stroke in SSSI

The precise mechanisms linking higher HCT levels with a lower risk of progressive stroke in SSSI remain incompletely understood. However, several plausible pathways could explain this association. One possible explanation is that higher HCT levels enhance the blood’s oxygen-carrying capacity, thereby improving oxygen delivery to the ischemic penumbra and limiting infarct growth.^14^ Enhanced oxygen delivery could help sustain the viability of at-risk brain tissue and reduce the extent of ischemic damage.

Additionally, higher HCT levels may indicate better hydration status and reduced hemodilution, both of which have been associated with favorable outcomes in acute ischemic stroke. Optimal hydration and lower levels of hemodilution help maintain blood viscosity and flow, thereby enhancing cerebral perfusion in ischemic regions.^21^ In contrast, anemia, which often correlates with lower HCT levels, can exacerbate oxidative stress and inflammatory responses in the brain, leading to increased production of reactive oxygen species (ROS). This oxidative stress can contribute to neuronal injury and activate inflammatory pathways, further worsening brain damage.^20^ The observed interaction between HCT and sensory deficits, specifically for NIHSS worsening by ≥4 points, suggests that the protective effect of higher HCT may be particularly relevant in patients with severe sensory dysfunction. This interaction may reflect the greater susceptibility of extensive or strategically located infarcts to the detrimental effects of low HCT levels on oxygen delivery and tissue preservation.

In summary, the potential mechanisms underlying the protective effects of higher HCT levels in SSSI may include improved oxygen delivery, better hydration status, reduced oxidative stress, and an interaction with specific neurological deficits such as sensory dysfunction. Future research should aim to further elucidate these mechanisms and explore how they can be leveraged to improve clinical outcomes in SSSI and other stroke subtypes.

### Clinical Implications and Future Directions

Our findings suggest that maintaining HCT levels within the normal range could be a viable strategy to reduce the risk of progressive stroke in SSSI patients, particularly those with severe sensory deficits. This strategy may involve avoiding excessive hemodilution and correcting anemia in the acute phase of stroke, potentially through interventions such as intravenous iron therapy. Excessive hemodilution decreases the concentration of red blood cells, thereby reducing the blood’s oxygen-carrying capacity and potentially worsening ischemic conditions. Conversely, correcting anemia can enhance oxygen delivery to ischemic brain tissue, potentially mitigating brain injury.

However, the optimal HCT target range remains to be established. While our study indicates that higher HCT levels are associated with a reduced risk of progressive stroke, it is essential to balance the benefits with the potential risks. Excessively elevated HCT levels could increase blood viscosity, impair microcirculation, and exacerbate ischemic conditions. Therefore, interventions aimed at modulating HCT levels should be carefully evaluated in randomized controlled trials to determine safe and effective targets for stroke patients.

Additionally, these results underscore the importance of regular monitoring of HCT levels in SSSI patients, particularly those with severe sensory deficits. Routine monitoring can aid in risk stratification and guide clinical decision-making. For instance, patients with lower HCT levels and severe sensory deficits may require more frequent monitoring and proactive preventive measures to mitigate the risk of neurological deterioration. Implementing standardized protocols for routine HCT monitoring in stroke care settings could help identify patients at higher risk and tailor interventions accordingly.

Future research should address several key areas. First, it is crucial to establish the optimal HCT range for preventing progressive stroke in SSSI patients through well-designed clinical trials. Second, investigating the mechanisms by which HCT influences stroke progression could provide deeper insights into potential therapeutic targets. Third, exploring genetic factors and individual patient characteristics that modulate the impact of HCT on stroke outcomes could enable the development of personalized treatment approaches. Finally, validating the association between lower HCT levels and increased early mortality risk in SSSI patients and elucidating the underlying mechanisms could guide targeted interventions to improve survival in this population.

### Strengths and Limitations

This study has several notable strengths. First, it utilized data from a large, nationwide, prospective registry with standardized data collection and outcome assessments, ensuring the reliability and generalizability of the findings. CNSR-III provided a robust dataset with comprehensive information on a large cohort of SSSI patients. Second, we focused on a specific subtype of ischemic stroke with a well-defined neuroimaging criterion, enhancing the specificity and relevance of our results to this patient population. Third, we accounted for a broad range of potential confounders, including demographic, clinical, and laboratory variables, to minimize the impact of confounding factors and provide accurate estimates of the association between HCT levels and progressive stroke. Fourth, we explored a potential dose-response relationship between HCT levels and progressive stroke risk and examined the interaction effects between HCT and patient characteristics, offering a more comprehensive understanding of HCT’s role in SSSI outcomes.

However, our study has limitations. First, HCT levels were only measured upon admission, and we did not assess the impact of changes in HCT levels during hospitalization on outcomes. Future studies should consider serial measurements of HCT to evaluate the dynamic relationship between HCT and progressive stroke. Second, we lacked detailed information on the etiology of SSSI, such as small artery occlusion or branch atheromatous disease, which may have different pathophysiologies and prognoses. Understanding these etiological subtypes could provide more nuanced insights into the role of HCT in different SSSI contexts. Third, while our findings suggest that higher HCT levels within the normal range may be beneficial, the optimal HCT target for preventing progressive stroke in SSSI remains to be determined. Further research is needed to identify an HCT range that balances the benefits of improved oxygen delivery with the potential risks of increased viscosity and thrombosis. Fourth, the observational design of this study precludes establishing causal relationships. Although we adjusted for numerous potential confounders, residual confounding by unmeasured variables cannot be ruled out. Future randomized studies are needed to confirm the causal role of HCT in influencing SSSI outcomes and to evaluate interventions that modulate HCT levels, such as phlebotomy, hydration strategies, or erythropoiesis-stimulating agents. Lastly, this study was conducted in a Chinese population, and the generalizability of our findings to other populations needs to be verified. Differences in genetic backgrounds, environmental factors, and healthcare systems may affect the relationship between HCT and stroke outcomes. Therefore, validation in diverse populations is necessary to ensure broader applicability.

### Implications of results for clinicians

This study demonstrates that higher admission HCT levels are independently associated with a reduced risk of progressive stroke in patients with SSSI, indicating a potential dose-response relationship and a significant interaction effect with severe sensory deficits. These findings suggest that maintaining HCT levels within an optimal range could be a viable strategy to reduce the risk of neurological deterioration, particularly in patients with severe sensory dysfunction. Additionally, lower HCT levels were independently associated with an increased risk of early mortality, underscoring the prognostic value of HCT in SSSI patients.

However, the observational design of our study and limitations such as the single-time-point measurement of HCT and the lack of detailed etiological data emphasize the need for further research. Future studies should focus on establishing optimal HCT targets, exploring the underlying mechanisms of HCT’s protective effects, and validating these findings in diverse populations through randomized controlled trials. Addressing these gaps could enhance our understanding of HCT’s role in stroke progression and inform the development of targeted interventions to improve outcomes for SSSI patients.

## Data Availability

All data are available upon reasonable request.

## Appendices

1. Appendix Table 1: Multivariate Association Between Hematocrit (HCT) Levels and Change in NIHSS Score (ΔNIHSS ≥ 1 Point)
2. Appendix Table 2: Multivariate Association Between Hematocrit (HCT) Levels and Change in NIHSS Score (ΔNIHSS ≥ 2 Points)
3. Appendix Table 3: Multivariate Analysis of Hematocrit (HCT) Levels and the Risk of Neurological Deterioration (ΔNIHSS ≥ 3 Points)
4. Appendix Table 4: Multivariate Analysis of Hematocrit (HCT) Levels and the Risk of Neurological Deterioration (ΔNIHSS ≥ 4 Points)
5. Appendix Table 5: Multivariate Analysis of Hematocrit (HCT) Levels and Changes in NIHSS Scores Across Motor and Sensory Subgroups
6. Appendix Table 6: Interaction Effects Between Hematocrit (HCT) Levels and Other Variables on Neurological Deterioration (ΔNIHSS ≥ 1 Point)
7. Appendix Table 7: Interaction Effects Between Hematocrit (HCT) Levels and Other Variables on Neurological Deterioration (ΔNIHSS ≥ 2 Points)
8. Appendix Table 8: Interaction Effects Between Hematocrit (HCT) Levels and Other Variables on Neurological Deterioration (ΔNIHSS ≥ 3 Points)
9. Appendix Table 9: Interaction Effects Between Hematocrit (HCT) Levels and Other Variables on Neurological Deterioration (ΔNIHSS ≥ 4 Points)

## Reference

1. Virani SS, Alonso A, Benjamin EJ, Bittencourt MS, Callaway CW, Carson AP, Chamberlain AM, Chang AR, Cheng S, Delling FN, et al. Heart Disease and Stroke Statistics-2020 Update: A Report From the American Heart Association. Circulation. 2020;141:e139–e596. doi: 10.1161/cir.0000000000000757

2. Sudlow CL, Warlow CP. Comparable studies of the incidence of stroke and its pathological types: results from an international collaboration. International Stroke Incidence Collaboration. Stroke. 1997;28:491–499. doi: 10.1161/01.str.28.3.491

3. Norrving B. Long-term prognosis after lacunar infarction. Lancet Neurol. 2003;2:238–245. doi: 10.1016/s1474-4422(03)00352-1

4. Jin D, Yang J, Zhu H, Wu Y, Liu H, Wang Q, Zhang X, Dong Y, Luo B, Shan Y, et al. Risk factors for early neurologic deterioration in single small subcortical infarction without carrier artery stenosis: predictors at the early stage. BMC Neurol. 2023;23:83. doi: 10.1186/s12883-023-03128-3

5. Arboix A, Martí-Vilalta JL. Lacunar stroke. Expert Rev Neurother. 2009;9:179–196. doi: 10.1586/14737175.9.2.179

6. Staaf G, Lindgren A, Norrving B. Pure motor stroke from presumed lacunar infarct: long-term prognosis for survival and risk of recurrent stroke. Stroke. 2001;32:2592–2596. doi: 10.1161/hs1101.098355

7. Wardlaw JM, Chabriat H, de Leeuw FE, Debette S, Dichgans M, Doubal F, Jokinen H, Katsanos AH, Ornello R, Pantoni L, et al. European stroke organisation (ESO) guideline on cerebral small vessel disease, part 2, lacunar ischaemic stroke. Eur Stroke J. 2024;9:5–68. doi: 10.1177/23969873231219416

8. Jeong HG, Kim BJ, Yang MH, Han MK, Bae HJ. Neuroimaging markers for early neurologic deterioration in single small subcortical infarction. Stroke. 2015;46:687–691. doi: 10.1161/strokeaha.114.007466

9. Yamamoto Y, Nagakane Y, Makino M, Ohara T, Koizumi T, Makita N, Akiguchi I. Aggressive antiplatelet treatment for acute branch atheromatous disease type infarcts: a 12-year prospective study. Int J Stroke. 2014;9:E8. doi: 10.1111/ijs.12200

10. In: Walker HK, Hall WD, Hurst JW, eds. Clinical Methods: The History, Physical, and Laboratory Examinations. Boston; 1990.

11. Diamond PT, Gale SD, Evans BA. Relationship of initial hematocrit level to discharge destination and resource utilization after ischemic stroke: a pilot study. Arch Phys Med Rehabil. 2003;84:964–967. doi: 10.1016/s0003-9993(03)00009-1

12. Furlan JC, Fang J, Silver FL. Acute ischemic stroke and abnormal blood hemoglobin concentration. Acta Neurol Scand. 2016;134:123–130. doi: 10.1111/ane.12521

13. Gotoh S, Hata J, Ninomiya T, Hirakawa Y, Nagata M, Mukai N, Fukuhara M, Ikeda F, Ago T, Kitazono T, et al. Hematocrit and the risk of cardiovascular disease in a Japanese community: The Hisayama Study. Atherosclerosis. 2015;242:199–204. doi: 10.1016/j.atherosclerosis.2015.07.014

14. Kimberly WT, Wu O, Arsava EM, Garg P, Ji R, Vangel M, Singhal AB, Ay H, Sorensen AG. Lower hemoglobin correlates with larger stroke volumes in acute ischemic stroke. Cerebrovasc Dis Extra. 2011;1:44–53. doi: 10.1159/000328219

15. Zhang R, Xu Q, Wang A, Jiang Y, Meng X, Zhou M, Wang Y, Liu G. Hemoglobin Concentration and Clinical Outcomes After Acute Ischemic Stroke or Transient Ischemic Attack. J Am Heart Assoc. 2021;10:e022547. doi: 10.1161/JAHA.121.022547

16. Wang Y, Jing J, Meng X, Pan Y, Wang Y, Zhao X, Lin J, Li W, Jiang Y, Li Z, et al. The Third China National Stroke Registry (CNSR-III) for patients with acute ischaemic stroke or transient ischaemic attack: design, rationale and baseline patient characteristics. Stroke Vasc Neurol. 2019;4:158–164. doi: 10.1136/svn-2019-000242

17. Wang Y, Li Z, Wang Y, Zhao X, Liu L, Yang X, Wang C, Gu H, Zhang F, Wang C, et al. Chinese Stroke Center Alliance: a national effort to improve healthcare quality for acute stroke and transient ischaemic attack: rationale, design and preliminary findings. Stroke Vasc Neurol. 2018;3:256–262. doi: 10.1136/svn-2018-000154

18. Cho AH, Kang DW, Kwon SU, Kim JS. Is 15 mm size criterion for lacunar infarction still valid? A study on strictly subcortical middle cerebral artery territory infarction using diffusion-weighted MRI. Cerebrovasc Dis. 2007;23:14–19. doi: 10.1159/000095753

19. Regenhardt RW, Das AS, Lo EH, Caplan LR. Advances in Understanding the Pathophysiology of Lacunar Stroke: A Review. JAMA Neurol. 2018;75:1273–1281. doi: 10.1001/jamaneurol.2018.1073

20. Sico JJ, Concato J, Wells CK, Lo AC, Nadeau SE, Williams LS, Peixoto AJ, Gorman M, Boice JL, Bravata DM. Anemia is associated with poor outcomes in patients with less severe ischemic stroke. J Stroke Cerebrovasc Dis. 2013;22:271–278. doi: 10.1016/j.jstrokecerebrovasdis.2011.09.003

21. Kaiafa G, Savopoulos C, Kanellos I, Mylonas KS, Tsikalakis G, Tegos T, Kakaletsis N, Hatzitolios AI. Anemia and stroke: Where do we stand? Acta Neurol Scand. 2017;135:596–602. doi: 10.1111/ane.12657

